# A novel benchmark for COVID-19 pandemic testing effectiveness enables the accurate prediction of new Intensive Care Unit admissions

**DOI:** 10.1101/2021.02.20.21252138

**Authors:** Dimitris Nikoloudis, Dimitrios Kountouras, Asimina Hiona

**Affiliations:** Center for Preventive Medicine & Longevity, Bioiatriki Healthcare Group, 11525 Athens, Greece

## Abstract

The positivity rate of testing is currently used both as a benchmark of testing adequacy and for assessing the evolution of the COVID-19 pandemic. However, since the former is a prerequisite for the latter, its interpretation is often conflicting. We propose as a benchmark for COVID-19 testing effectiveness a new metric, termed ‘Severity Detection Rate’ (SDR), that represents the daily needs for new Intensive Care Unit (ICU) admissions, per 100 cases detected (t-i) days ago, per 10,000 tests performed (t-i) days ago. Based on the announced COVID-19 monitoring data in Greece from May 2020 until August 2021, we show that beyond a certain threshold of daily tests, SDR reaches a plateau of very low variability that begins to reflect testing adequacy. Due to the stabilization of SDR, it was possible to predict with great accuracy the daily needs for new ICU admissions, 12 days ahead of each testing data point, over a period of 10 months, with Pearson r = 0.98 (p = 10^−197^), RMSE = 7,16. We strongly believe that this metric will help guide the timely decisions of both scientists and government officials to tackle pandemic spread and prevent ICU overload by setting effective testing requirements for accurate pandemic monitoring. We propose further study of this novel metric with data from more countries to confirm the validity of the current findings.

## Introduction

Although no country knows at any point the true total number of COVID-19 cases, it is crucial for public health administrations to be confident that the daily testing performed is stably representative of that number. Effective testing provides health professionals and officials with a clear picture of SARS-CoV-2 spread within the community, as well as of the dynamics of COVID-19 pathology, and guides them for prompt and adequate interventions towards containment of the pandemic at the local and national levels.

The percentage of tests that return a positive result, also known as the “positivity rate”, is an important outcome of testing that is used both as a benchmark for testing adequacy and as a metric for assessing the current spread of the virus^1,2^. However, this dual usage presents an inherent drawback in entrusting the metric in any one of two possible ways: is a high positivity rate due to a high number of infected individuals, or due to a low number of tests performed? A rule of thumb says that a positivity rate of 5% is too high, and the WHO suggests that the positivity rate should rest below that threshold for a length of at least two weeks before officials decide to progressively reopen professional and social activities^1^. Another evidence-based perception suggests that the positivity rate must remain below 3% to ensure that surveillance is broad and accurate enough^2^. However, these rules may only cover either the virus spread surveillance criterion or that of testing adequacy, not both. Indeed, officials often respond to a high positivity rate both with an increase in testing and with measures to restrict virus transmission, such as social distancing and soft or hard lockdowns. But by doing so, it is expectedly hard to timely assess the true rate of the virus spreading out, or being contained, as the new higher levels of testing must be stabilized for a length of time before allowing again to reliably follow the pandemic dynamics. In such a scenario, if health officials rely only on the positivity rate metric, the timing of the response would lag and thus be almost invariably suboptimal.

Fundamentally, a metric that would serve as a benchmark for the effectiveness of COVID-19 testing should not concurrently be used for assessing the evolution of the pandemic, as the former is a prerequisite for the latter and therefore the interpretation would be conflicting; indeed, the health administrations of a country should be confident that a sufficient number of tests is performed to effectively track the virus spread. However, if such a metric also implemented measurable outcomes of the pandemic in the community (e.g., number of deaths, number of ICU admissions, etc.), they could introduce by their more factual nature a link between expectation and actuality, since the outcomes of COVID-19 are inherently tied to the virus’s pathogenesis. Therefore, such a link could, in theory, introduce a benchmarkable step of convergence towards a soft cap (threshold) that would in turn reflect testing adequacy, *e*.*g*., usually a maximized or minimized value, or a state of minimized variation. In this report, we present an easy-to-implement metric that we developed while independently monitoring and analyzing COVID-19 pandemic evolution in Greece, which considers outcomes that are already monitored in most countries, such as the daily numbers of human losses, COVID-19 patients in the ICU (Intensive Care Units), and patients who are being discharged from the ICU. In our example we show that this metric displays remarkable output stability when a certain threshold of daily testing is reached, which to our view clearly reflects testing adequacy. Furthermore, we validated its benchmarking efficiency by forecasting, not only with high accuracy but also great precision, the total daily needs for new ICU admissions, roughly two weeks in advance, over a period of 10 months.

## Methods

The national monitoring data for the evolution of the COVID-19 pandemic in Greece were retrieved from the Hellenic National Public Health Organization^3^ and Greek Government’s official daily announcements^4^. Specifically, the daily official announcements included the following parameters: (a) number of new COVID-19 cases detected, (b) number of deaths due to COVID-19, (c) total number of COVID-19 ICU patients, (d) total number of COVID-19 patients discharged from ICU, (e) total number of SARS-CoV-2 PCR tests performed^5^, and (f) total number of SARS-CoV-2 rapid antigen tests performed^6^.

Based on the available data, we defined *the daily needs for new COVID-19 ICU admissions* as number U:

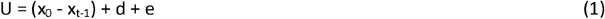

where:

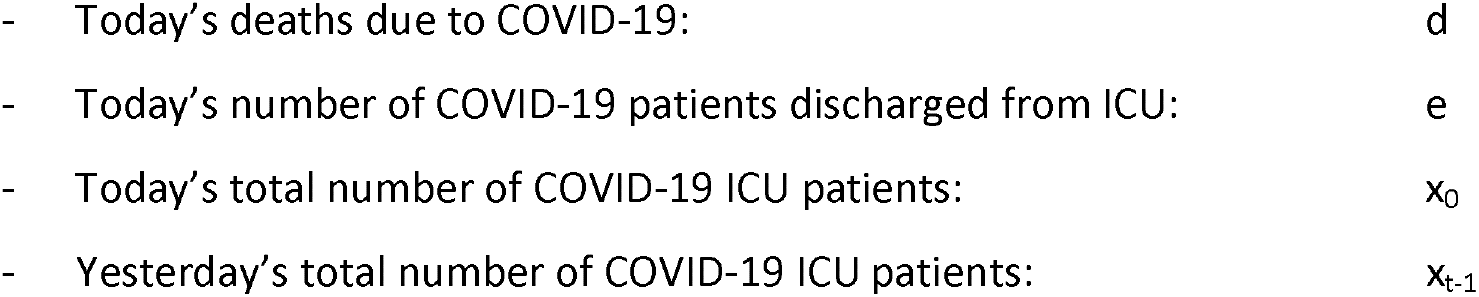

This number U represents the actual daily new COVID-19 ICU admissions, plus those patients who died in the community (not in ICU), whom we theorize to have required ICU admission, hence the definition of *the daily needs for new COVID-19 ICU admissions*.

Next, we defined as the Severity Detection Rate with a time lag (t-i) (SDR_i_), a metric that represents the percentage of patients who require ICU admission, per new cases, detected (t-i) days ago, per 10,000 tests, performed (t-i) days ago:

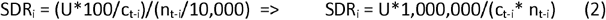

where:

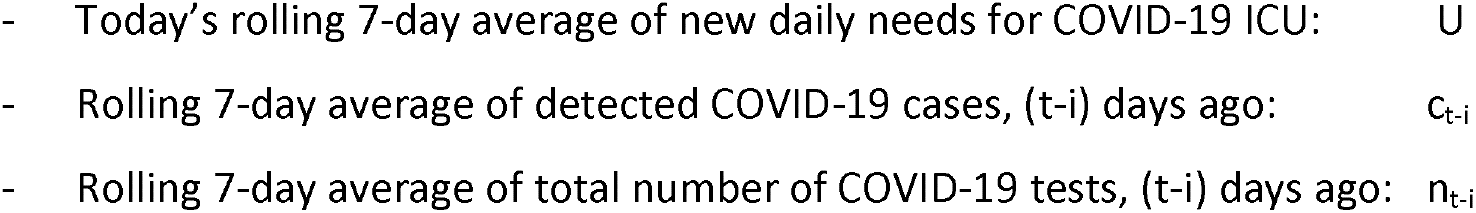

Tests in Greece were performed freely by any individual who wanted to get tested, in selected hospitals, or in most private diagnostic centers and clinics, or in mobile testing hubs, dispatched by the public healthcare administration. Also, an individual may get tested in regular intervals (*e*.*g*., up to twice per week), as requested by their employer or the administration, due to the nature of their profession. To the best of our knowledge, only one swab is taken from the individual per test, in Greece. Furthermore, the reported COVID-19 cases detected, and daily tests performed, are used for the official calculation of positivity rate, announced routinely by the country’s healthcare administration^3^; if multiple tests per individual were simply added to the total daily number, this would constitute a systematic error in the calculation of positivity rate. Therefore, for the reasons explained above, for this analysis, the daily number of tests reported publicly is presumed to represent unique individuals.

For initial data exploration, the lag of Severity Detection Rate (SDR) metrics was set to 14 days, which means that the current day’s critical outcomes of COVID-19 (*i*.*e*., ICU admission or death in the community) were attributed to COVID-19 cases detected 14 days ago. For the identification of the optimal lag point between the critical outcomes of COVID-19 and the detected cases, we searched within an interval between 7 to 21 days, in the period 17/10/20 to 31/1/21 of the dataset, for the most stable correlation between the numerator (number U) and the denominators ([cases_t-i_ * tests_t-i_]) of the metrics studied. The best correlation was obtained for a lag of 12 days (i=12) (see Discussion section) and therefore, for consistency, all charts and tables reflect this optimal time lag (i=12).

Finally, for completeness of the study, we also defined as ICU admission Rate with a time lag (t-i) (henceforth “ICU Rate”, IR_i_), a metric that represents the percentage of patients who require ICU admission, per new cases, detected (t-i) days ago:

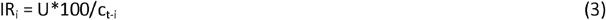

where:

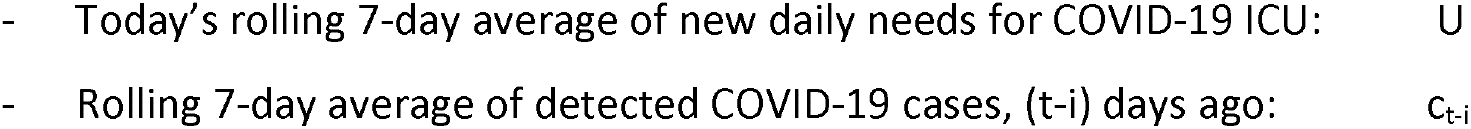

IR metric is essentially a simpler form of the SDR metric, which doesn’t take into account the number of daily tests performed. As we wanted to also evaluate its predictive performance, we doubled every piece of analysis performed on the SDR metric, on the IR metric as well. The related charts and tables are not part of the Results section in favor of clarity for the main metric presented (SDR), but are, nonetheless, commented upon in the Discussion section.

The dataset was locked on August 8^th^ 2021.

## Results

For observation, the daily evolution of SDR_12_, from the 7 ^th^ of May 2020 onwards, was traced on the same chart versus the observed number of daily ICU needs, the positivity rate and the corresponding number of testing samples (Figure 1).

**Figure 1.**
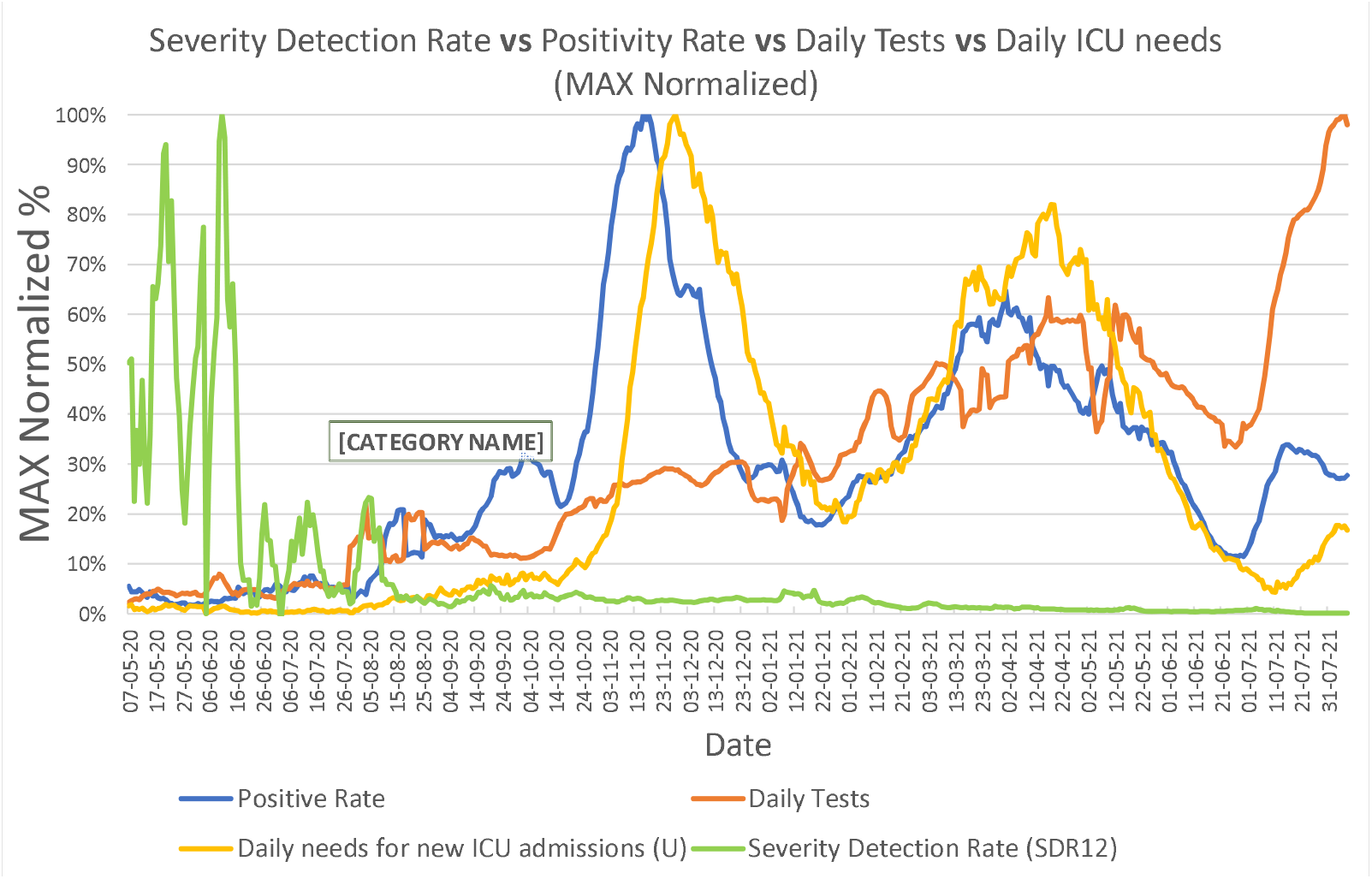
Comparison of trendlines of Severity Detection Rate, Daily needs for new ICU admissions, Positivity rate, and number of Daily Tests, in the period from 7/5/2020 to 8/8/2021. The Daily needs for new ICU admissions and the number of Daily Tests represent rolling 7-day averages. Severity Detection Rates and Positivity Rates were calculated from the rolling 7-day averages of their components. All numbers were normalized by their maximum value in the examined period.

Compared to the other quantities, the SDR metric shows a remarkable stabilization past the time mark on approximately 20/8/2020, which also corresponds to the attainment of an average daily testing number of 10,000/day. From that point forward, the observed daily ICU needs, the positivity rate and the testing rate continue to fluctuate independently and considerably, but without accordingly perturbing SDR stabilization.

The rate of daily testing in Greece has been scaled up significantly on four occasions, approximately (a) on 29/7/2020, (b) on 17/10/2020, (c) on 1/2/2021 and (d) on 11/7/2021. As the new testing levels were preserved after each scale-up, it is possible to define 5 distinct periods of testing intensity thus far during the COVID-19 pandemic in Greece. Interestingly, a sixth distinct period is noted between 1/5/2021 and 10/7/2021, where, inversely, a steady reduction in the number of daily tests is observed, although the daily average number of tests is preserved from the immediately previous period. We qualified this behavior as noteworthy and chose to study the respective period separately. We therefore characterized the SDR number and the rates of testing for each of the following time intervals: (i) 1/5/2020 - 28/7/2020, (ii) 29/7/2020 – 16/10/2020, (iii) 17/10/2020 – 31/1/2021, (iv), 1/2/2021 – 30/4/2021, (v) 1/5/2021 – 10/7/2021, and (vi) 11/7/2021 – 8/8/2021 (Table 1).

**Table 1.** Characterization of the Severity Detection Rate and the number of daily tests for each of the six time intervals of distinct testing levels in Greece.

| Distinct periods of testing levels |  |  |  |  |  |  |
| --- | --- | --- | --- | --- | --- | --- |
| intervals | 1/5/2020 - 28/7/2020 |  | 29/7/2020 – 16/10/2020 |  | 17/10/2020 - 31/1/2021 |  |
|  | SDR<sub>12</sub> | Samples | SDR<sub>12</sub> | Samples | SDR<sub>12</sub> | Samples |
| max | 92.0% | 7309 | 7.0% | 20310 | 3.2% | 31602 |
| average | 20.1% | 4051 | 2.7% | 12861 | 2.1% | 24377 |
| median | 14.3% | 3992 | 2.6% | 12453 | 2.0% | 24743 |
| min | 0.1% | 1400 | 1.1% | 9706 | 1.3% | 17315 |
| sd | 19.6% | 1260 | 1.0% | 2439 | 0.4% | 3172 |
| cv | 0.97 | 0.31 | 0.36 | 0.19 | 0.19 | 0.13 |
| intervals | 1/2/2021 - 30/4/2021 |  | 1/5/2021 - 10/7/2021 |  | 11/7/2021 - 27/7/2021 |  |
|  | SDR<sub>12</sub> | Samples | SDR<sub>12</sub> | Samples | SDR<sub>12</sub> | Samples |
| max | 1.6% | 58578 | 1.0% | 57206 | 0.12% | 76262 |
| average | 0.9% | 42905 | 0.4% | 42150 | 0.11% | 69974 |
| median | 0.9% | 41906 | 0.4% | 41922 | 0.11% | 73005 |
| min | 0.5% | 29504 | 0.2% | 30871 | 0.10% | 58583 |
| sd | 0.3% | 7510 | 0.2% | 7190 | 0.01% | 5919 |
| cv | 0.32 | 0.18 | 0.36 | 0.17 | 0.06 | 0.08 |

Tripling the average daily rate of testing (from 4K to 12K) in the second (ii) interval brought a 7-fold lower average value of SDR (20.1% / 2.7% ∼ 7.4), with a remarkable 20-fold decrease (19.6% / 1%) in the Standard Deviation (SD) of SDR, and a concomitant 3-fold decrease in the CV (Coefficient of Variation) of SDR (0.97/0.36 ∼ 2.7). Further doubling of the average daily number of tests (from 12 K to 24 K) in the third (iii) interval again brought an equivalent decrease in the SDR SD (1.0% / 0.4% = 2.5) although the average value of SDR was now only moderately diminished by approximately 30% (2.7% / 2.1% ∼ 1.29), indicating a tendency towards stabilization of the SDR value and a continuous reduction of the Standard Deviation (SD). Overall, it is noteworthy that specifically the average and SD values of SDR continued to drop consistently in all 6 periods.

We then traced the values of SDR metric against the daily number of tests. The SDR values display a strong correlation with the daily number of tests, employing power regression (Spearman r = -0.90, p = *10^−167^*, N = 451) and suggest that beyond a threshold of daily tests performed, SDR becomes significantly stabilized (Figure 2); for Greece, this stabilization begins once the number of daily tests exceeds the mark of 10,000 per day.

**Figure 2.**
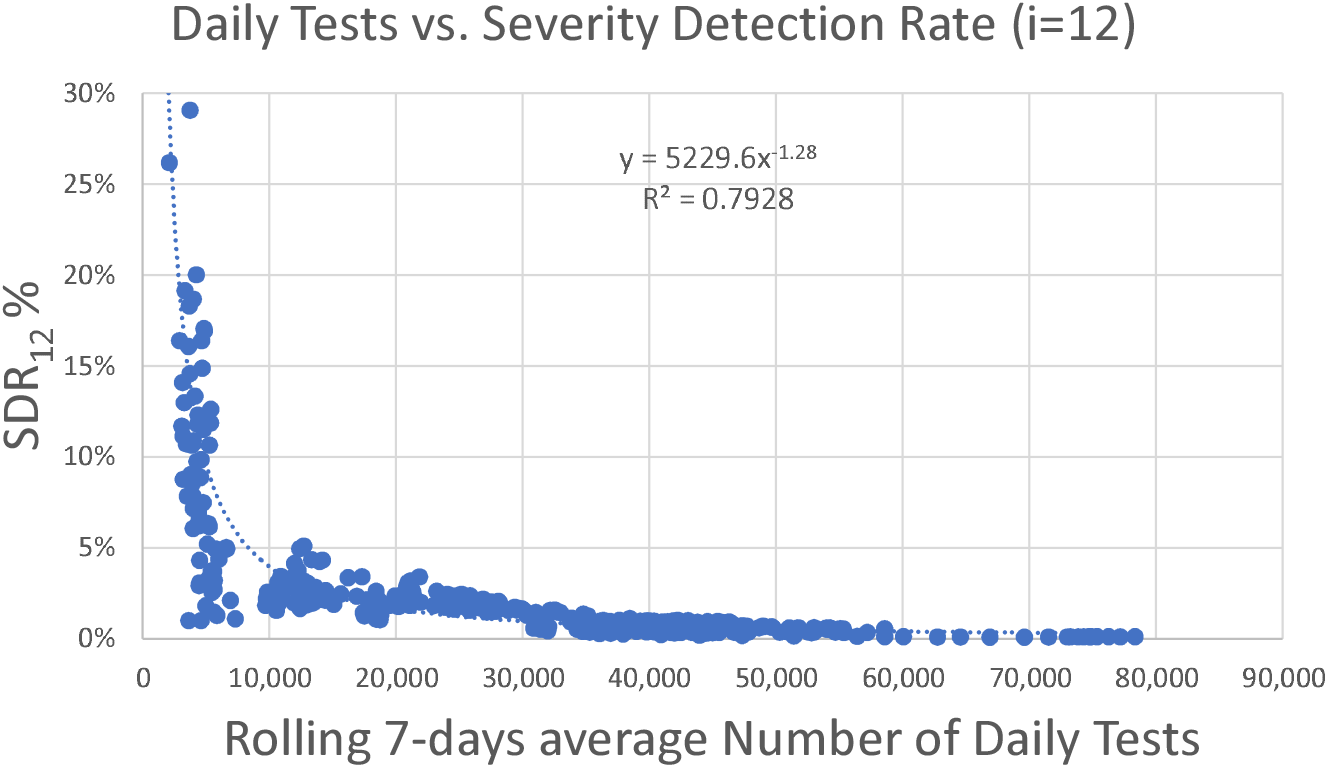
Correlation between the Severity Detection Rate and rolling 7-day averages of the number of daily tests, with Spearman r = -0.90, p = 10^−167^, N = 451. Numbers of daily tests derived from the period from 15/5/2020 to 8/8/2021.

The next step was to study the correlation between the numerator (number U) and the denominator ([cases_t-i_ * tests_t-I_]) of SDR metric, for the period 17/10/2020 to 8/8/2021 (Figure 3). The starting period was chosen to be the same as the start of testing period (iii) (Table 1). Before that date, both the numbers of new daily needs for ICU and daily cases were relatively low (Figure 1) and therefore of smaller interest to the specific study, *i*.*e*., when added to the rest of the data, the respective correlation is innately stronger due to the near-baseline nature of the data points prior to 17/10/2020.

**Figure 3.**
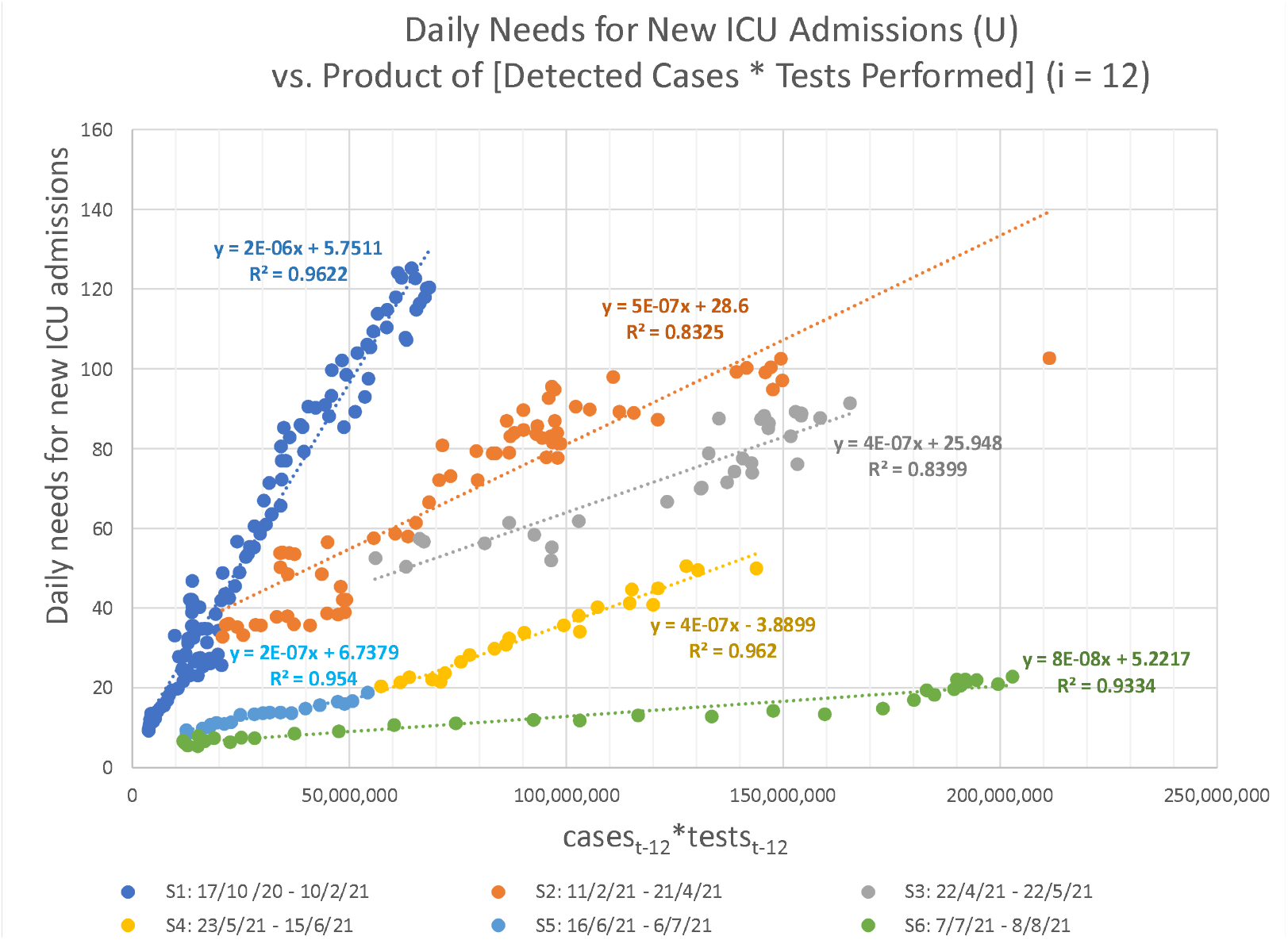
Correlation of the numerator and denominator of SDR, i.e., number U versus the product (cases_t-12_* tests_t-12_), with a lag of 12 days. Numbers of daily tests and detected cases were derived from the period from 17/10/2020 to 8/8/2021.

Finally, we applied the linear regression equations to forecast the rolling 7-day average daily needs for new ICU admissions, 12 days ahead of each data point of daily announced cases and tests, for the corresponding periods, *i*.*e*., from 17/10/2020 to 8/8/2021. The forecast employing the SDR regression equations (Figure 3) proved very accurate (Pearson r = 0.98, p = 10^−197^, RMSE = 7.16; with n = 296, observed U[max] = 125, U[average] = 51) (Figure 4). Expectedly, as can be noticed in Figure 4B, most of the few intense discrepancies in the fitted values are observed around dates of transition from one regression equation to another; a rolling regression window could possibly help improve the forecast of even these phases. Overall, forecasting with the use of Severity Detection Rate proved to be functional as it indicated a very strong agreement between the predicted and observed values for a period of nearly 10 months, which included the two major pandemic waves in the country, thus far.

**Figure 4.**
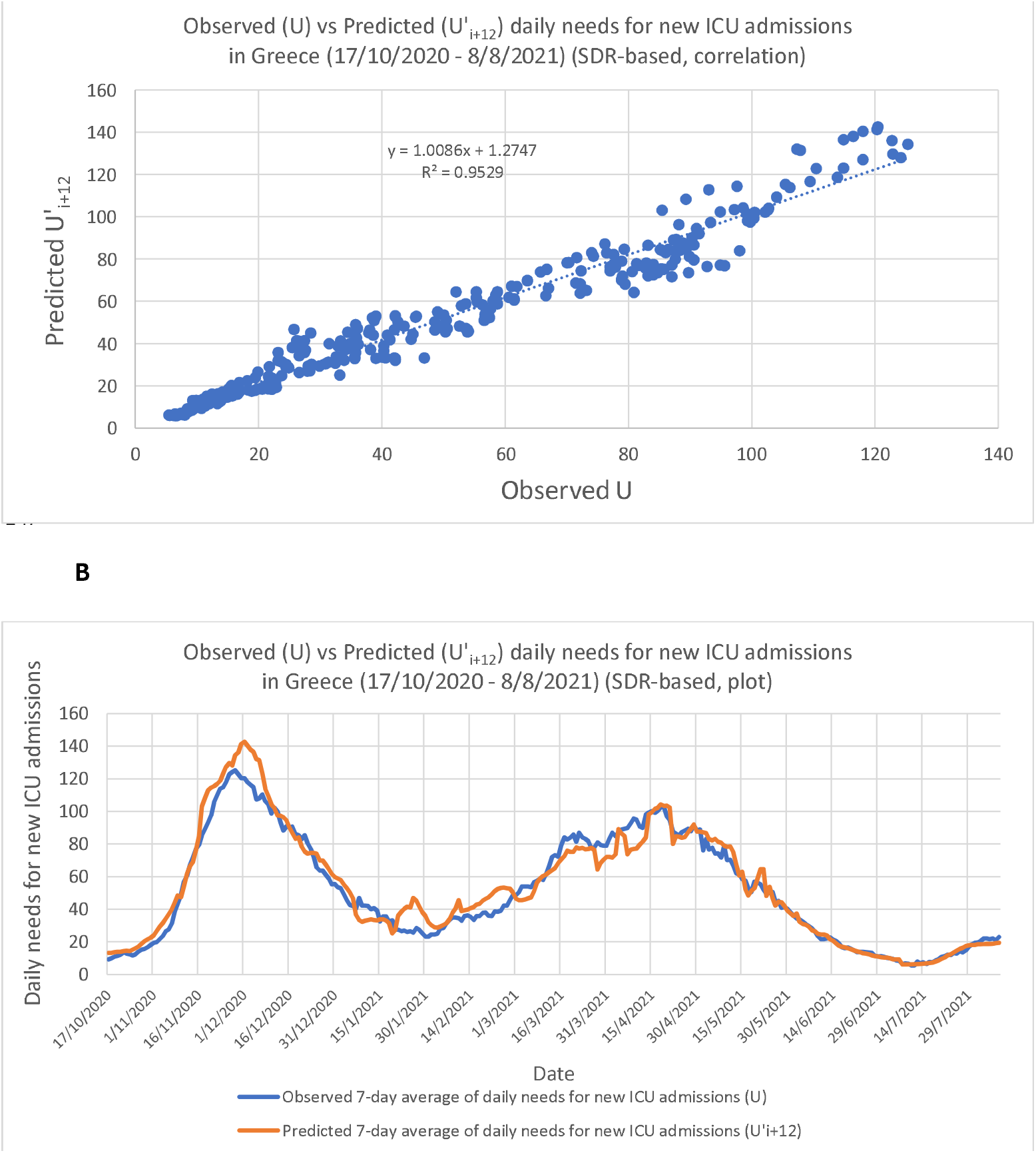
(A) Correlation between observed and predicted daily needs for new ICU admissions for the period between 17/10/2020 and 8/8/2021 employing Severity Detection Rate regression equations, with Pearson r = 0.98, p = 10^−197^, RMSE = 7.16; with n = 296, observed U[max] = 125, U[average] = 51. (B) The respective time-series plots for visual inspection of the fit.

## Discussion

We have shown that beyond a threshold of daily tests performed, SDR reaches a plateau that displays very low variation. This threshold appears roughly around the 10,000 daily samples mark in Greece, a country of approximately 11 million people, but this number is expected to vary greatly from country to country depending on the total population, rural density, societal particularities, population immune profile, and sampling strategies^7^. Reaching that threshold should not mean that there is no need for further increase in the number of daily tests, as it is strongly suggestive that the more tests a country performs, the more informative the results are about the actual viral spread in the community, and consequently health administrations are in better position to respond accordingly. In terms of the SDR metric, more daily tests appear to further decrease its variation (Table 1). The weaker its variation, the stronger the correlation coefficient between *the numerator and denominator of SDR, i*.*e*., *number U versus the product* (*casest-i* testst-i*), *and therefore*, the more accurately we can predict the number of daily needs for new ICU admissions, t+i days in advance. In the studied example, predictions were highly accurate with an average daily number of tests as high as 24,000 (Table 1), which resulted in a SD of the SDR of 0.4%. As the SD of the SDR showed a consistent decrease over a period of 15 months in our studied example (Table 1), we propose it can possibly act as an actual numerical threshold that denotes the attainment of the SDR plateau.

As a direct consequence of this potential predictability, when SDR establishes a plateau, we consider that the bulk of daily tests is returning a set of positive cases that is stably representative of the current spread of the virus. Therefore, the SDR metric constitutes a benchmark of testing effectiveness. The metric is potentially efficient at a local level as well, if cases that require delocalization, *e*.*g*., due to lack of available ICU locally, are effectively tracked and taken into account. As the full segmentation of the necessary data was not available at a local level for the present study, it was not possible to evaluate the effects of viral spread uniformity across the country and, more specifically, the metric’s behavior due to disproportionate testing intensities locally, e.g., higher number of tests in districts with lower viral load, and relatively lower numbers of daily tests in districts with higher true viral load. In such a case, it would be helpful to apply the SDR monitoring at a local level.

The metric’s median value is expected to decrease monotonically and with decreasing variation as daily tests increase, or due to the gradual containment of the virus, immunization of the population, thanks to an efficient vaccination program, improvement of therapeutic protocols that reduce the number of very severe cases, or even a significant reduction in the average age of infected individuals due to the efficient protection of the elderly. Conversely, the metric’s median value may increase (interrupting the plateau) if the viral spread becomes greatly enhanced with time, *e*.*g*., due to the prevalence of a new more infectious variant^8,9,10^, without the testing levels catching up. In such a case the SDR’s median will increase disproportionately and beyond its expected variability.

In order to comprehend the nontrivial nature of the plateau attainment and retainment in the plot of SDR versus the number of daily tests (Figure 2), it is useful to look more carefully at some notable boundaries of the SDR metric. For instance, if it was possible to test the entire population every day for newly infected individuals (minus the individuals that are already known to be infected), then the “discovery” of every new infection case would be guaranteed (assuming 100% accurate tests). With a number of daily tests as big as the entire population and with the highest possible number of detected cases (*i*.*e*., equal to the actual cases), the SDR value becomes [U / ((actual new cases) * population)] with the denominator assuming its greatest possible value, hence producing the lowest possible SDR.

In a different approach that hypothetically guarantees the detection of all the actual new infected cases (without testing the entire population), we can consider testing all the newly infected individuals, and only them, so that the number of daily tests becomes equals to the number of new infections (again, assuming 100% testing accuracy). In this case the SDR value becomes [U/(actual cases * actual cases) = U/ actual cases^2^]. Whether the possible values of the SDR metric can be bigger or smaller than the value obtained in this second hypothetical scenario, depends on whether the product [cases_t-i_ * tests_t-i_] is smaller or bigger than the square of the number of actual new infection cases (see mathematical demonstration, below). Finally, as the theoretical maximum of all the possible SDR values we may consider the case where the denominator [cases_t-i_ * tests_t-i_] is equal to 1, and therefore SDR would be equal to number U. Specifically:

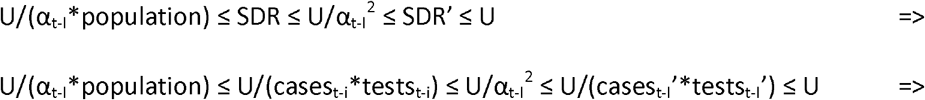

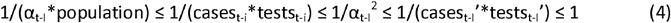

therefore:

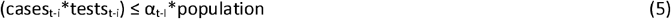

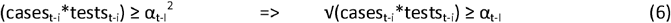

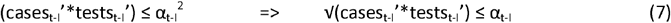

where:

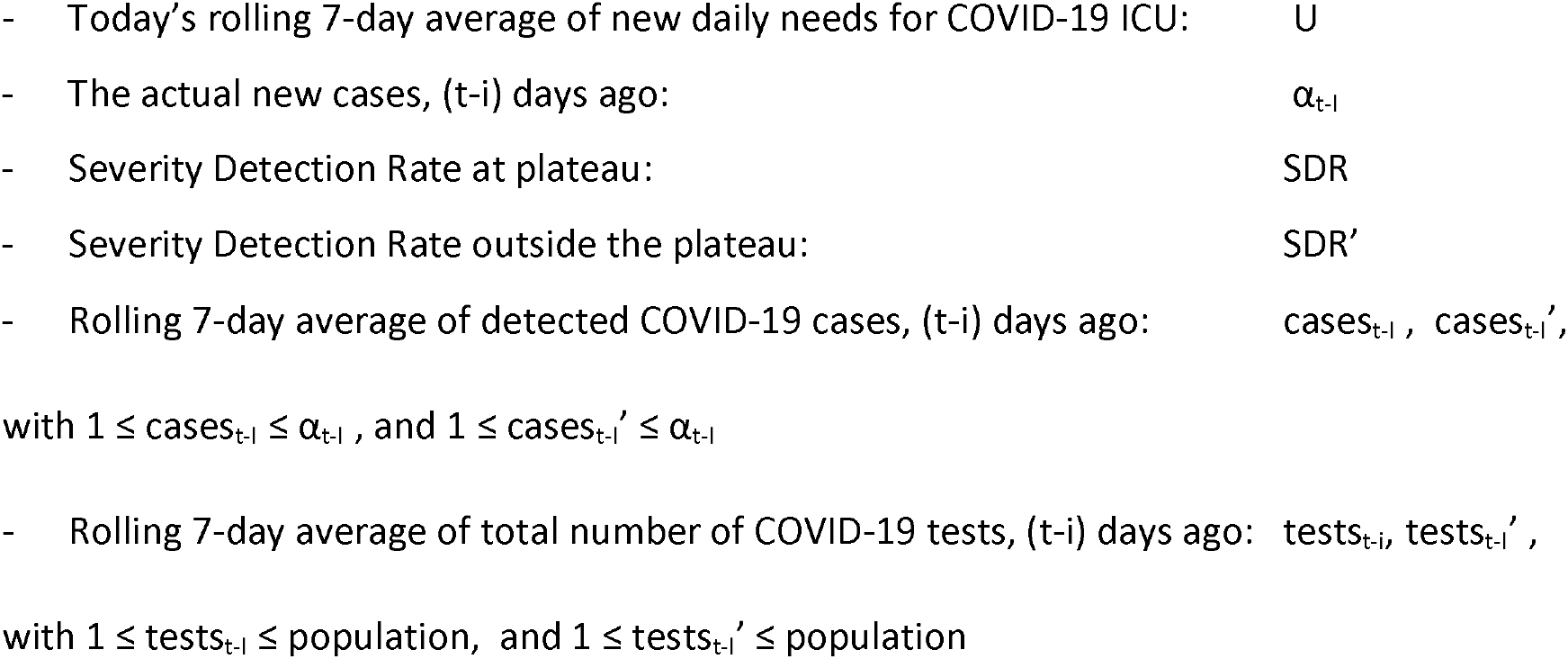

Inequality (5) is trivial as the number of actual new cases (α_t-I_) and the entire population of the country, or area of interest, are by definition the highest possible values of the product (cases_t-i_*tests_t-i_). However, inequality (6) describes a situation where the number of tests can only be equal or greater than α_t-i_^2^/cases_t-i_, and which may increase up to the number of the entire population, causing the reduction of the SDR value till its described minimum of U/(α_t-I_*population). Inequality (7), inversely, describes a situation where the number of tests can only be equal or lower than α t-I ^2^/cases t-I ‘, and which may decrease to as low as 1 test, causing the increase of the SDR value to its maximum that equals the number U.

Therefore, because of this demonstrated relationship between the number of daily tests and the number of actual new infections, we theorize that in a plot of SDR versus the number of daily tests, the observed plateau is a consequence of the SDR starting to adopt values that are smaller than U/α^2^. Inversely we observe values outside the plateau as long as SDR adopts values greater than U/α^2^. This is potentially what happened around the mark of 10,000 tests in our studied example (roughly around 20/8/2020), with the product (cases * tests) increasing almost 10-fold within a few days and presumably becoming greater than the square of the actual new cases, thus collapsing the SDR variability into the observed plateau (Figures 2 & 5). The importance of the plateau being, as previously explained, the reduction of the metric’s variability (i.e., Standard Deviation), enabling a correspondingly robust forecasting of ICU needs, (t+i) days ahead of each datapoint.

**Figure 5.**
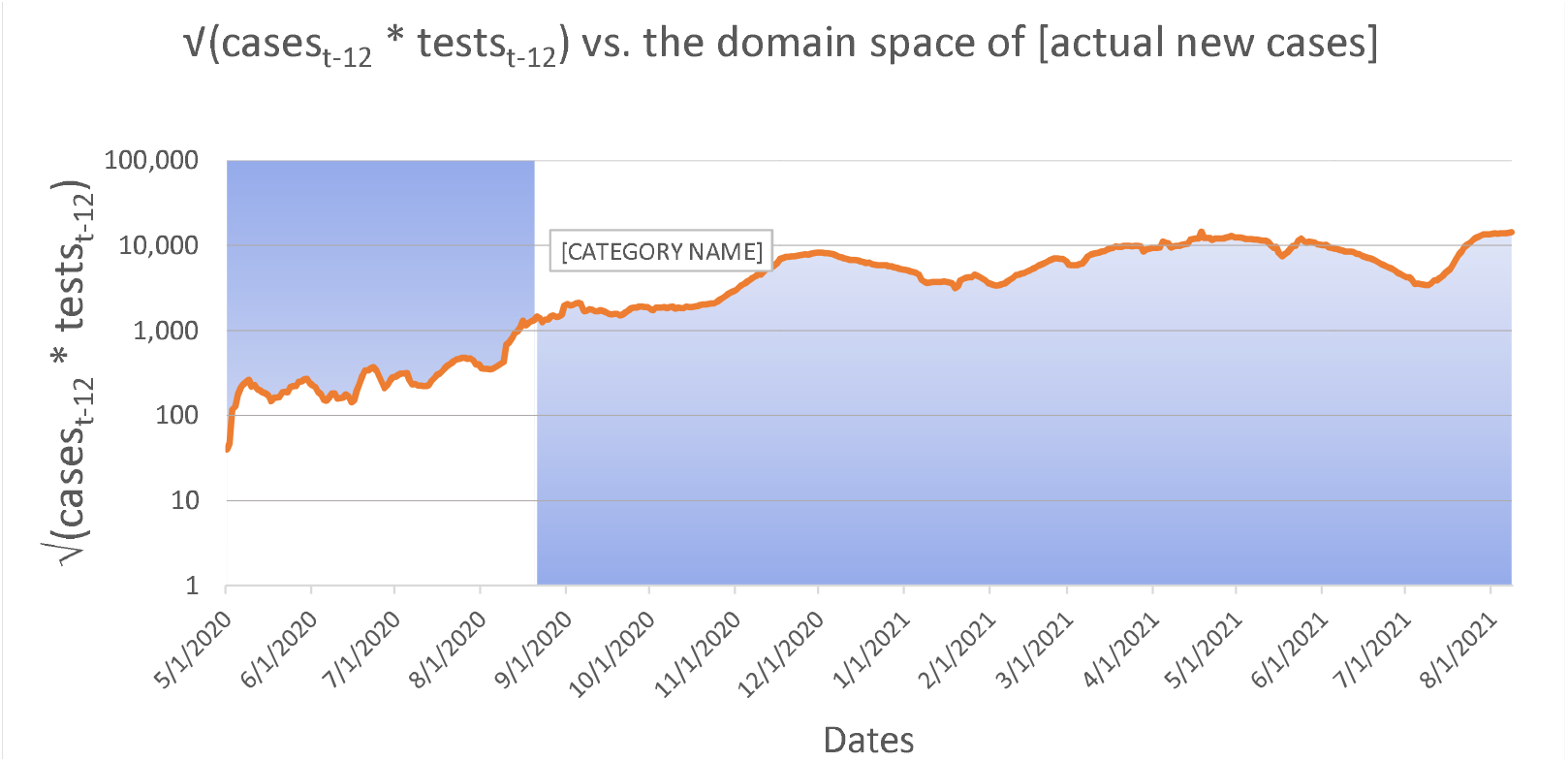
Visualization of the domain space where the number of actual new cases are to be found hypothetically (blue area), relatively to the square root of the product [cases_t-12_ * tests_t-12_] (orange line), before and after the date of 8/20/2020, which marked the beginning of the SDR plateau.

In the context of the regression analysis of the daily needs for new ICU admissions (U) vs. the product of [Detected Cases * Performed Tests] (Figure 3), significant changes in the SDR median would be reflected as changes in the slope and/or the intercept of the regression line. Specifically, changes in the slope most likely translate into two possibilities: (A) a change in virulence (*i*.*e*., how many individuals per group of 100 positive cases, per 10,000 tests, are expected to develop very severe COVID-19, given a theoretical zero regression intercept), or (B) a modification in sampling parameters (*e*.*g*., testing more or fewer asymptomatic persons, or testing a younger subset of the population). Accordingly, a change in the intercept will likely signify either (a) changes in viral prevalence^7,11^, as the intercept represents a fixed number U for a theoretical x=0, (*i*.*e*., a number of individuals with very severe COVID-19, while no cases are detected), or (b) changes in testing accuracy^7,11^, with intercept values closer to zero reflecting optimal accuracy. Rolling 3-weeks regression windows could be employed to detect dynamic changes of the pandemic. The study of all the available confounding factors (*e*.*g*., prevalence of new virus variant, changes in sampling strategies, changes in testing parameters, characteristics of areas infected, lockdown and other measures’ status, ages of tested and infected individuals, etc.) is required to discern which exact change is responsible for the observed new disease dynamics, and the SDR derived regression analysis can provide significant hints as to the direction of the change. In any of the above cases, an important shift of the SDR would signify an important change in the pandemic parameters, which in turn would dictate a specific course of action for the authorities, appropriate for each case.

In Table 2 we contrast the regression parameters (*i*.*e*., slope, intercept and R^2^) against important factors of the ongoing pandemic, such as, Delta variant prevalence, vaccination levels, and lockdown periods. What is most notable is the stable slope decrease of the regression equations, over all 6 periods examined, which is compatible with a decrease in population-level severity/virulence. This is to be expected, given the long periods of the applied lockdown measures and the ongoing mass vaccination program in the country (reaching 50% population coverage of fully vaccinated individuals on 8/8/2021). As presented in the previous paragraph, another factor that can possibly lower the SDR slope is a significant change in sampling parameters, in a way where the group of asymptomatic individuals that are being tested becomes considerably increased, a situation that results inherently to fewer detected cases than the group of symptomatic individuals. While it is hard to discern the potential contribution of each factor with just the publicly available data, it is, nonetheless, possible to calculate a 9.5-fold total drop in the observed severity between the beginning and ending of the six periods (17/10/2020 -> 8/8/2021), after adjusting for the obvious contribution of the change in the average number of cases and tests (Table 2):

**Table 2.**
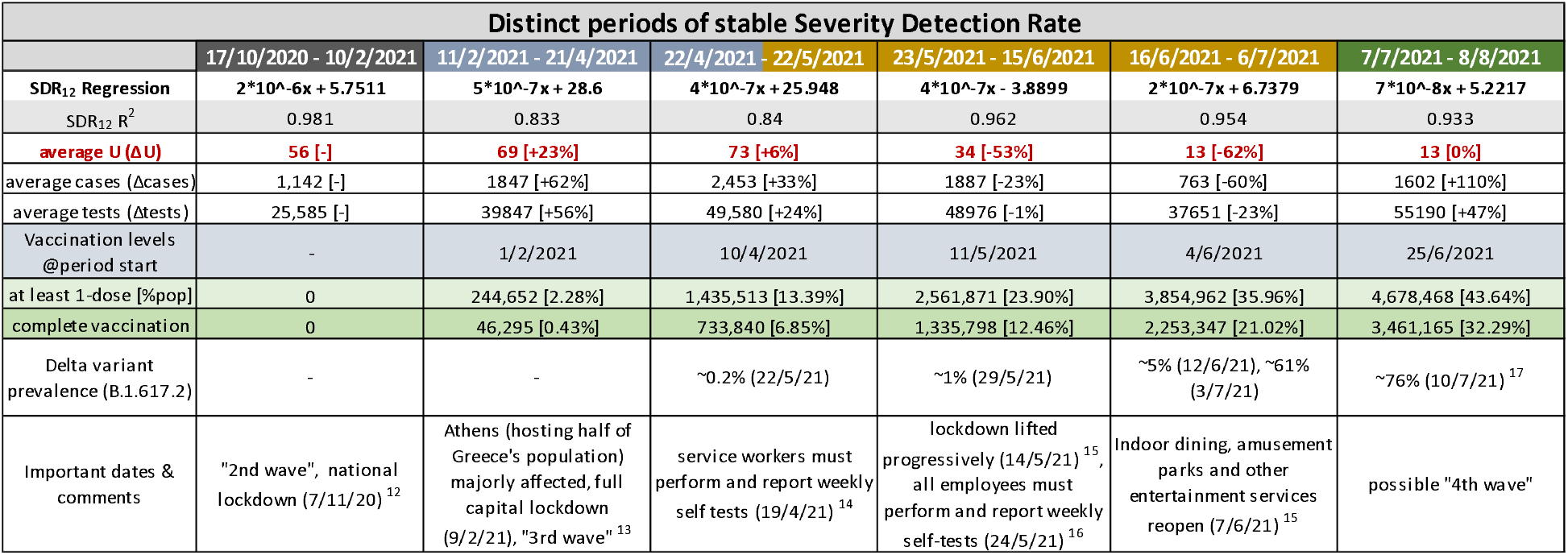
Regression equations per distinct period of stable Severity Detection Rate, with respective average numbers of observed daily needs for new ICU admissions, cases detected and tests performed, vaccination coverage at the beginning of each period, percent of Delta variant prevalence in cases detected, along with important dates and comments that potentially influenced the course of the pandemic in Greece (each period is color-coded with reference to the respective ‘distinct periods of testing levels’, in Table 1).

| Distinct periods of stable Severity Detection Rate |  |  |  |  |  |  |
| --- | --- | --- | --- | --- | --- | --- |
|  | 17/10/2020 - 10/2/2021 | 11/2/2021 - 21/4/2021 | 22/4/2021 - 22/5/2021 | 23/5/2021 - 15/6/2021 | 16/6/2021 - 6/7/2021 | 7/7/2021 - 8/8/2021 |
| SDR<sub>12</sub> Regression | $2 \cdot 10^{-6}x + 5.7511$ | $5 \cdot 10^{-7}x + 28.6$ | $4 \cdot 10^{-7}x + 25.948$ | $4 \cdot 10^{-7}x - 3.8899$ | $2 \cdot 10^{-7}x + 6.7379$ | $7 \cdot 10^{-8}x + 5.2217$ |
| SDR<sub>12</sub> R<sup>2</sup> | 0.981 | 0.833 | 0.84 | 0.962 | 0.954 | 0.933 |
| average U (ΔU) | 56 [-] | 69 [+23%] | 73 [+6%] | 34 [-53%] | 13 [-62%] | 13 [0%] |
| average cases (Δcases) | 1,142 [-] | 1847 [+62%] | 2,453 [+33%] | 1887 [-23%] | 763 [-60%] | 1602 [+110%] |
| average tests (Δtests) | 25,585 [-] | 39847 [+56%] | 49,580 [+24%] | 48976 [-1%] | 37651 [-23%] | 55190 [+47%] |
| Vaccination levels @period start | - | 1/2/2021 | 10/4/2021 | 11/5/2021 | 4/6/2021 | 25/6/2021 |
| at least 1-dose [%pop] | 0 | 244,652 [2.28%] | 1,435,513 [13.39%] | 2,561,871 [23.90%] | 3,854,962 [35.96%] | 4,678,468 [43.64%] |
| complete vaccination | 0 | 46,295 [0.43%] | 733,840 [6.85%] | 1,335,798 [12.46%] | 2,253,347 [21.02%] | 3,461,165 [32.29%] |
| Delta variant prevalence (B.1.617.2) | - | - | ~0.2% (22/5/21) | ~1% (29/5/21) | ~5% (12/6/21), ~61% (3/7/21) | ~76% (10/7/21)<sup>17</sup> |
| Important dates & comments | "2nd wave", national lockdown (7/11/20)<sup>12</sup> | Athens (hosting half of Greece's population) majorly affected, full capital lockdown (9/2/21), "3rd wave"<sup>13</sup> | service workers must perform and report weekly self tests (19/4/21)<sup>14</sup> | lockdown lifted progressively (14/5/21)<sup>15</sup>, all employees must perform and report weekly self-tests (24/5/21)<sup>16</sup> | Indoor dining, amusement parks and other entertainment services reopen (7/6/21)<sup>15</sup> | possible "4th wave" |

Δ(slope, 17/10/2020 -> 8/8/2021): (2*10^−6^ / 7*10^−8^) = 28.6 - Unadjusted fold change in severity

Δ(average SDR denominator, 17/10/2020 -> 8/8/2021): (1,602*55,190) / (1,142*25,585) = 3 - fold change in product (cases * tests)

Δ(observed severity, 17/10/2020 -> 8/8/2021): 28.6 / 3 = 9.5 - Adjusted fold change in severity

On the contrary, the intercept oscillates considerably between periods, ranging from +28.6 to - 3.9. As explained previously, increases of the intercept may be attributed to greater viral spread in the community, as was the case in the second period (11/2/2021 – 21/4/2021), when Athens, the capital, saw a great increase in infected cases, which signaled the beginning of the 3^rd^ wave of the pandemic in Greece. Besides viral spread, the other factor that influences the intercept is the accuracy of the tests performed, i.e., potential false positives and false negatives, due to poor test specificity, test sensitivity, or yet undetectable levels of the virus in asymptomatic infected individuals who simply got tested too early in the course of the disease. Regarding Delta variant prevalence (B.1.617.2), representing 90% of cases in Greece on 8/8/2021, it doesn’t appear to be affecting the severity of the disease (*i*.*e*., a slope increase), however it is possibly contributing to the intercept increase from 16/6/2021 onwards, with its greater transmissibility potential, as reported by other studies^18^. Overall, the slope and intercept of SDR-based regression equations offer an additional layer of information, which, in conjunction with other metrics and parameters, may create a better understanding of the pandemic’s dynamics.

We called this new metric Severity Detection Rate, as its representation of the percentage of very severe COVID-19 outcomes is modulated by the number of tests performed. It is essentially a standardization of the very severe cases ratio over the infected individuals, with the rate of daily testing. In other words, the Severity Detection Rate becomes representative of the proportion of people who need ICUs out of the total cases once a sufficient threshold of daily testing rate (hence ‘detection rate’) is achieved.

As presented in the Methods section, for a more complete examination, we also defined the percentage of patients who require ICU admission, per new cases detected (t-i) days ago, as ICU Rate (IR). If, in theory, the total number of tests became equal to the entire population of a country (or the area of interest), then the SDR metric would be the same as the IR metric, as the ‘number of tests’ parameter would be removed from the denominator (as redundant), and both would practically represent the true percentage of critical patients per infected individual. In order to assess the predictive potential of the IR metric, we have repeated for IR every piece of analysis that was performed on the SDR metric throughout this study.

Regarding forecasting, the conclusion drawn by this parallel analysis is that the IR metric performed as well as the SDR metric, in the analyzed example (Figures 3-S, 4-S, Table 2-S). On top of this, the IR metric would probably have the advantage of simplicity when communicated in the general public, as it represents a more comprehensible concept: the number of very severe cases per infected individuals. We therefore believe that the IR metric may be used in cases where the population-level COVID-19 testing surveillance of the pandemic is well established, by efficient and sufficient testing. Nonetheless, we support that by including the number of daily tests performed, the SDR metric is inherently more suitable for a wider range of surveillance scenarios, *e*.*g*., when the testing strategies and pandemic parameters (e.g., number & type of tests, geographical/ occupational/ age targeting, contact tracing efficacy, transmissibility of the virus, etc.) are more volatile in time. In different countries, or in specific areas of interest, it is still possible for the IR-based monitoring to fail to return regression coefficients as strong as in our studied example. In those cases, it would be necessary to switch to SDR-based monitoring to ensure that a threshold of sufficient testing has been reached (*i*.*e*., plateau formation). In any case, although more studied examples are required to better understand the potential practical differences between the two metrics, since they both showed equal forecasting performances, we believe that SDR is the more well-rounded metric, which can be efficiently used in potentially very diverse situations of pandemic surveillance.

## Conclusions

Taken together, the monitoring of the Severity Detection Rate and the forecasting of number U (*i*.*e*., daily needs for ICU) should be viewed as integral parts of the currently employed epidemiological toolbox, i.e., the positivity rate, efficient contact tracing for determination of the basic reproduction number R _0_ ^19,20^, and wastewater-based surveillance^21,22^. The metric introduces the goal for authorities to minimize its variation by means of a sufficient number of daily tests and an adequate sampling strategy. Once this goal is achieved, accurate forecasting of daily needs for new ICU admissions becomes possible. With accurate forecasting, number U becomes in essence a quantitative metric for the severity of the pandemic.

In Figure 6 we detail all the proposed steps for population-level surveillance of COVID-19 pandemic using the Severity Detection Rate metric. For monitoring SDR Standard Deviation, a minimum of 3-weeks rolling window interval is suggested empirically, as this interval includes the roughly 2-week lag period between case detection and ICU intubation. The recommended surveillance model provides three distinct advantages: (1) a measurable threshold for adequacy of tests performed, (2) important qualitative information regarding the current dynamics of the pandemic (virulence, prevalence, testing accuracy, etc.) that are reflected by changes in the slope/intercept of the regression analysis, and (3) the ability to accurately predict the ICU needs, t+I days ahead.

**Figure 6.**
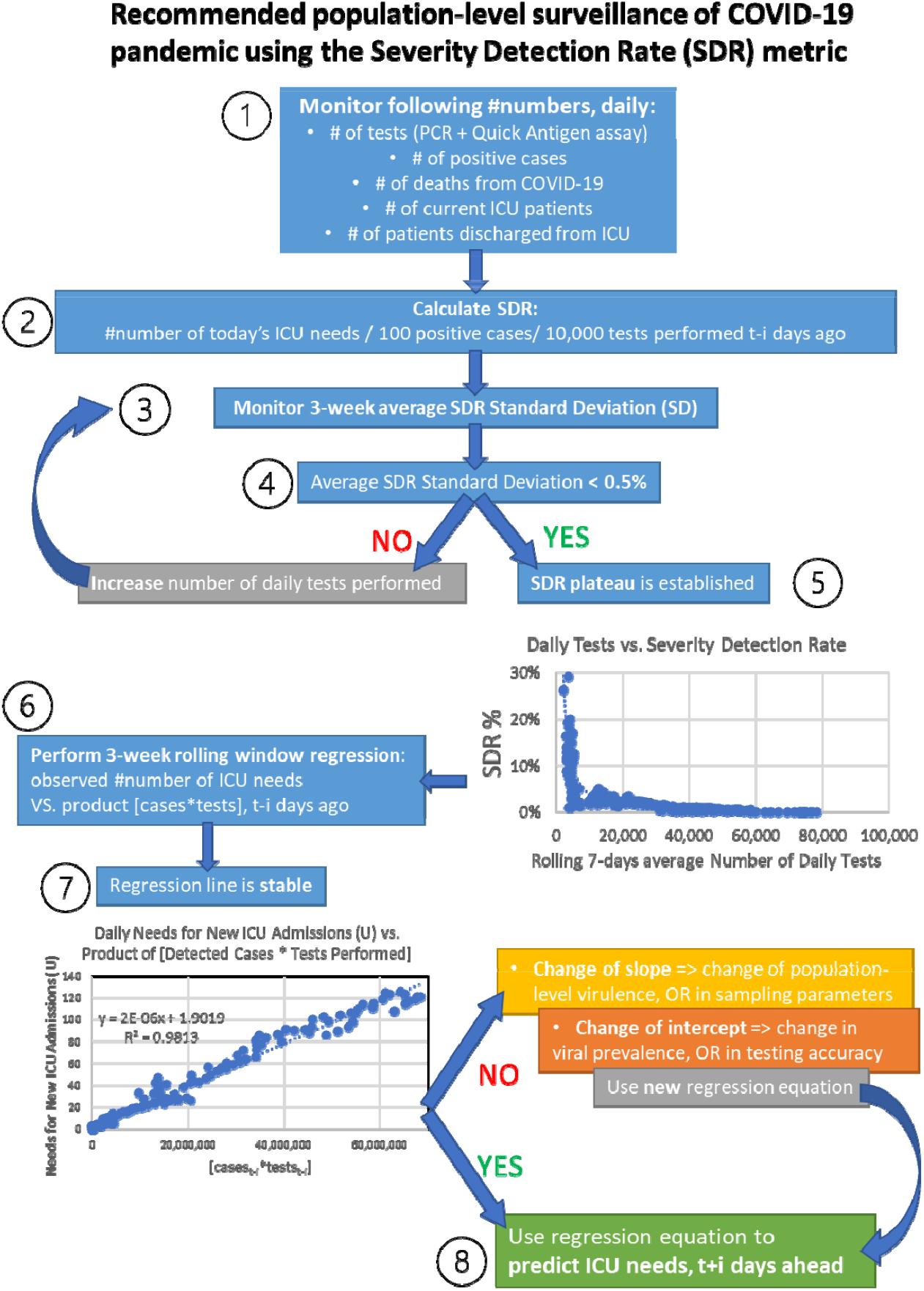
Stepwise schema detailing the logic and requirements for population-level surveillance of COVID-19 pandemic with the use of the Severity Detection Rate metric.

We strongly believe that the explicit tracking of this novel metric enhances the visibility of viral spread and dynamics and may procure an accurate outlook of the upcoming needs for ICU admissions well in advance, which should serve as an early warning system for COVID-19 health establishments and resources. We therefore suggest further study of Severity Detection Rate with data from more countries, as well as at a local level wherever possible, to confirm the proposed functionality and utility of this metric.

## Data Availability

All primary data used are publicly available.

https://eody.gov.gr/epidimiologika-statistika-dedomena/ektheseis-covid-19/

https://invite.viber.com/?g2=AQAVNiDVlfjrIEtaYLf1s2sUzRrpfLVlfLVg4J8wkdNKMUSnUcQWJxnXH0Os1heH&lang=el

## Notes

### Competing Interest Statement

All authors are employed by Bioiatriki Healthcare Group

### Funding Statement

No specific funding was received for this study.
All authors are employed by Bioiatriki Healthcare Group.

### Author Declarations

No IRB/oversight body approvals were required for this study.

### Summary of Updates

Update to match final peer reviewed version.

## References

1. Dowdy D, D’Souza G. COVID-19 Testing: Understanding the “Percent Positive”. Johns Hopkins Bloomberg School of Public Health. Accessed 31 August 2021. Available from: https://www.jhsph.edu/covid-19/articles/covid-19-testing-understanding-the-percent-positive.html

2. Siddarth D, Katz R, Graeden E, Analytics T, Allen D, Tsai T. 2020. Evidence Roundup: Why positive test rates need to fall below 3%. Harvard Global Health Institute. Accessed 31 August 2021. Available from: https://globalhealth.harvard.edu/evidence-roundup-why-positive-test-rates-need-to-fall-below-3

3. Hellenic National Public Health Organization. Daily Reports COVID-19. Accessed 31 August 2021. Available from: https://eody.gov.gr/epidimiologika-statistika-dedomena/ektheseis-covid-19/

4. Greek Government’s official community on Viber network. Official COVID-19 update. Accessed 31 August 2021. Available from: https://invite.viber.com/?g2=AQAVNiDVlfjrIEtaYLf1s2sUzRrpfLVlfLVg4J8wkdNKMUSnUcQWJxnXH0Os1heH&lang=el

5. Corman VM, Landt O, Kaiser M, Molenkamp R, Meijer A, Chu DKW, Bleicker T, Brünink S, Schneider J, Schmidt ML, Mulders DGJC, Haagmans BL, van der Veer B, van den Brink S, Wijsman L, Goderski G, Romette J-L, Ellis J, Zambon M, Peiris M, Goossens H, Reusken C, Koopmans MPG, Drosten C. 2020. Detection of 2019 novel coronavirus (2019-nCoV) by real-time RT-PCR. Euro Surveill. 25(3):pii=2000045. doi.org: 10.2807/1560-7917.ES.2020.25.3.2000045.

6. Lambert-Niclot S, Cuffel A, Le Pape S, Vauloup-Fellous C, Morand-Joubert L, Roque-Afonso AM, Le Goff J, Delaugerre C. 2020. Evaluation of a Rapid Diagnostic Assay for Detection of SARS-CoV-2 Antigen in Nasopharyngeal Swabs. J Clin Microbiol. 23;58(8):e00977–20. doi: 10.1128/JCM.00977-20.

7. Mercer TR, Salit M. 2021. Testing at scale during the COVID-19 pandemic. Nat Rev Genet 22, 415–426. doi: 10.1038/s41576-021-00360-w

8. Elbe S, Buckland-Merrett G. 2017. Data, disease and diplomacy: GISAID’s innovative contribution to global health. Glob Chall. 10;1(1):33–46. doi: 10.1002/gch2.1018.

9. Hadfield J, Megill C, Bell SM, Huddleston J, Potter B, Callender C, Sagulenko P, Bedford T, Neher RA. 2018. Nextstrain: real-time tracking of pathogen evolution. Bioinformatics 1;34(23):4121–4123. doi: 10.1093/bioinformatics/bty407.

10. Forster P, Forster L, Renfrew C, Forster M. Phylogenetic network analysis of SARS-CoV-2 genomes. 2020. Proc Natl Acad Sci U S A. 28;117(17):9241–9243. doi: 10.1073/pnas.2004999117.

11. Nichols JD, Bogich TL, Howerton E, Bjørnstad ON, Borchering RK, Ferrari M, Haran M, Jewell J, Pepin KM, Probert WJM, Pulliam JRC, Runge MC, Tildesley M, Viboud C, Shea K. 2021. Strategic testing approaches for targeted disease monitoring can be used to inform pandemic decision-making. PLoS Biol 19(6): e3001307. doi: 10.1371/journal.pbio.3001307

12. Krinis N. 2020. Greece to Enter Lockdown to Fight Second Covid-19 Wave. Greek Travel Pages. Accessed 31 August 2021. Available from: https://news.gtp.gr/2020/11/05/greece-enter-lockdown-fight-second-covid-19-wave

13. Koutantou A. 2021. Greek premier orders full lockdown in Athens after surge in coronavirus cases. Reuters. Accessed 31 August 2021. Available from: https://www.reuters.com/world/greek-premier-orders-full-lockdown-athens-after-surge-coronavirus-cases-2021-02-09

14. Reuters Staff. 2021. Greece orders COVID self-testing for service workers. Reuters. Accessed 31 August 2021. Available from: https://www.reuters.com/article/health-coronavirus-greece-tests-idUSL8N2M740B

15. ESN COVID-19 Official Announcements & News. Timeline for the loosening of the lockdown measures in Greece. 2021. Erasmus Student Network Greece. Accessed 31 August 2021. Available from: https://esngreece.gr/covid-19-official-announcements-news

16. GTP editing team. 2021. All Employees in Greece Must Self-test for Covid-19. Greek Travel Pages. Accessed 31 August 2021. Available from: https://news.gtp.gr/2021/05/24/all-employees-greece-must-self-test-covid-19

17. European Centre for Disease Prevention and Control. SARS-CoV-2 variants of concern as of 26 August 2021. Accessed 31 August 2021. Available from: https://www.ecdc.europa.eu/en/covid-19/variants-concern

18. Centers for disease control and prevention. Delta Variant: What We Know About the Science. Accessed 31 August 2021. Available from: https://www.cdc.gov/coronavirus/2019-ncov/variants/delta-variant.html

19. Macdonald G (1952). The analysis of equilibrium in malaria. Tropical Diseases Bulletin. 49 (9): 813–829. ISSN 0041-3240. PMID 12995455.

20. Delamater PL, Street EJ, Leslie TF, Yang YT, Jacobsen KH. 2019. Complexity of the basic reproduction number (R0). Emerg. Infect. Dis. 25, 1–4.

21. Wu Y, Guo C, Tang L, Hong Z, Zhou J, Dong X, Yin H, Xiao Q, Tang Y, Qu X, Kuang L, Fang X, Mishra N, Lu J, Shan H, Jiang G, Huang X. 2020. Prolonged presence of SARS-CoV-2 viral RNA in faecal samples. Lancet Gastroenterol Hepatol. 5(5):434–435. doi: 10.1016/S2468-1253(20)30083-2. Epub 2020 Mar 20.

22. Polo D, Quintela-Baluja M, Corbishley A, Davey LJ, Andrew CS, David WG, Jesús LR. 2020. Making waves: Wastewater-based epidemiology for COVID-19 – approaches and challenges for surveillance and prediction. Water Research, 186, 116404. doi: 10.1016/j.watres.2020.116404.

